# Biomarker potential of vitreous microRNA in retinal disease: a meta-analysis

**DOI:** 10.1101/2024.03.25.24304858

**Authors:** Diana Joseph, Brian Grover, Michael Telias

**Affiliations:** Flaum Eye Institute, University of Rochester Medical Center, 601 Elmwood Ave, Rochester, NY 14642

**Keywords:** micro-RNA, vitreous humor, retina, liquid biopsy, biomarker, diagnosis, retinal disease

## Abstract

**Background:** Acquired retinal diseases such as proliferative diabetic retinopathy and age-related macular degeneration pose significant challenges in diagnosis and prognosis. The vitreous fluid, situated in the posterior chamber of the eye behind the lens, holds a close relationship with the inner retina. Within this milieu, retinal cells secrete a diverse array of biomolecules, potentially harboring vital biomarkers. Among these, short, non-coding micro-RNAs (miRNAs) emerge as promising candidates. Their dynamic regulation by various gene signaling mechanisms, enhanced resistance to degradation, and secretion via separate exocytotic pathways make them particularly significant. Alterations in vitreal miRNA profiles may reflect pathological states and offer insights into disease etiology and progression.

**Abstract:** We conducted a comprehensive meta-analysis of 22 peer-reviewed studies to assess the potential of vitreous miRNAs as biomarkers for retinal diseases. Our analysis demonstrates the potential utility of miRNAs as biomarkers in specific retinal pathologies. We show that miR-142, miR-9, and miR-21 emerge as robust biomarker candidates, displaying consistent and significant alterations correlating with proliferative vitreoretinal diseases. We also address the methodological challenges encountered in characterizing vitreous miRNA content, including the absence of standardized purification, amplification, and analysis protocols, as well as the scarcity of true control samples. Moreover, we make the case for the adoption of specific housekeeping genes and data normalization techniques to standardize miRNA analysis in the vitreous and explore potential methodologies for obtaining vitreous samples from healthy individuals.

**Conclusion:** Vitreous miRNAs hold promise as potential biomarkers for various retinal diseases, with miR-142, miR-9, and miR-21 emerging as particularly promising candidates. Enhancing methodologies for vitreous sampling and miRNA analysis presents an opportunity to expand the repertoire and utility of miRNA biomarkers in retinal disease diagnosis and prognosis.

**Graphic abstract:** 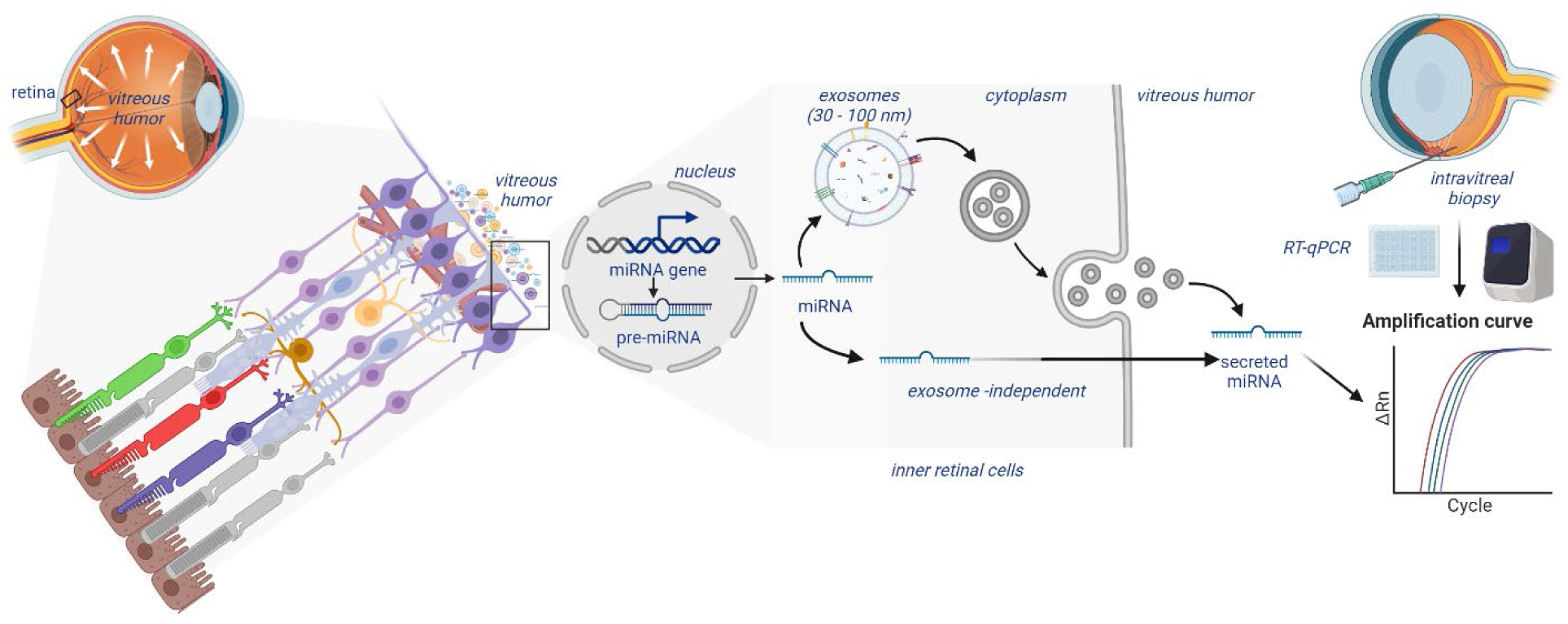

From left to right, the diagram shows the location of the retina and the vitreous humor within the eye; the microanatomy of the retinal layers showcasing the secretion of exosomes and biomolecules into the vitreous from its anterior side (inner retinal layers); a magnified illustration of miRNA secretion process from gene expression in the cell nucleus to exosome -dependent and - independent secretion pathways; and the process of intravitreal biopsy for collection and analysis of miRNA expression using quantitative PCR methods.

## Background

Retinal diseases can be hereditary, such as a retinitis pigmentosa, a disease affecting the photoreceptor layer of the retina, while others are acquired. Acquired diseases may be related to underlying cardiovascular disease (i.e. hypertensive retinopathy, retinal vein occlusion), diabetes mellitus (i.e. non-proliferative diabetic retinopathy -NPDR-, proliferative diabetic retinopathy -PDR-), or malignancy (i.e. melanoma, lymphoma). Other acquired retinal conditions are not directly caused by systemic disease but are more likely to occur in patients with retinal risk factors. For example, patients with pathological myopia (PM) and lattice degeneration are at greater risk for retinal detachment (RD) (1). Lack of known genetic causes behind acquired retinal diseases precludes strategies aimed to preventing their occurrence (e.g.: by preimplantation genetic screening), or to diagnose them before onset, as it can happen with late onset inherited retinal dystrophies.

Moreover, as compared to hereditary retinal diseases, acquired retinal diseases are far more prevalent in the population, and represent some of the most common causes behind vision loss and blindness. For example, age-related macular degeneration (AMD) has a prevalence of over 10% in US adults over the age of 40 and is the leading cause of blindness in elderly persons (2). Diabetic retinopathy (DR) affects over 40% of the growing population of patients with diabetes mellitus (3). Many of these diseases also progress over time. For example, both AMD and DR can progress to more severe forms, characterized by abnormal growth of blood vessels in the retina (4, 5), which further deteriorates vision in an irreversible way in the absence of treatment. As such, early detection of acquired retinal diseases is critical to preventing further vision loss.

However, early diagnosis of retinal diseases can be challenging. Exam skills, such as ophthalmoscopy, and imaging tools, like optical coherence tomography, enable visualization of the affected layers of the retina in most diagnoses. However, not all diagnoses are clear, and sometimes further investigation is required. For example, intraocular tumors can be mistaken for uveitis, necessitating a chorio-retinal biopsy to rule out malignancy (6). Not only do biopsies provide a more definitive diagnostic tool, but they can also further our basic scientific understanding of the molecular pathology in the sampled tissue. Unfortunately, chorio-retinal biopsies are invasive procedures, and the indications are limited to identifying infectious organisms affecting the retina, evaluating uveitis, and excluding malignancy. As a result, for most retinal diseases which do not necessitate a diagnostic biopsy, our understanding of the molecular pathogenesis is limited. Animal models, such as rodents, rabbits, dogs, and non-human primates, have undergone retinal biopsies for the study of retinal disease (7–9). However, the utility of animal models is limited by sample size for larger animals and differences in physiology and anatomy between smaller animals and humans. Moreover, recapitulating a non-inherited disease in an animal which does not develop the disease spontaneously, is complicated, time-consuming, and usually produces partial results, where the gap between humans and model animals remains large.

Liquid biopsy of the vitreous humor may provide a less invasive alternative to the chorio-retinal biopsy. The vitreous humor is a gel-like fluid filling the posterior chamber of the eye between the lens and the retina, in direct contact with the inner retina. The vitreous is composed of water, hyaluronan, hyalocytes (i.e. macrophages populating the posterior vitreous cortex), collagen and additional soluble proteins (10). The vitreous is important for maintaining the round structure of the eye and plays a role in refraction as light must pass through the vitreous to reach the retina (11). The vitreous humor is accessible for liquid biopsy (i.e.: vitrectomy) via an incision in the sclera. Analogous to how cerebrospinal fluid is collected through a lumbar puncture in order to evaluate central nervous system pathology, without the need to access the brain or spinal cord, vitreous humor can be biopsied to study basic and clinical aspects of retinal pathology (12). For example, in patients with PDR, retinal ischemia induces the release of vascular endothelial growth factor (VEGF) in the retina, which then leaks into the vitreous body, resulting in higher levels of vitreal VEGF (10, 13). In vitreoretinal diseases, such as PDR, proliferative vitreoretinopathy (PVR), and rhegmatogenous retinal detachment (RRD), the inflammatory proteins AAT, APOA4, ALB, and TF were found to be elevated compared to healthy controls, indicating that these proteins may contribute to their pathogenesis (14). Similarly, patients with diabetic macular edema, AMD, idiopathic epiretinal membranes (ERM), and macular telangiectasia type 2 show abnormal expression of various proteins in the vitreous humor (15). Furthermore, the stability created by interaction between hyaluronic acid and collagen fibrils within the vitreous also slows clearance, allowing persistence of molecules associated with disease (10). The variations in vitreous composition reflecting retinal pathology along with their persistence in the vitreous body suggest that the fluid may hold utility as a biomarker and diagnostic tool. In the future, vitreous humor may surpass chorio-retinal biopsy and provide insight into the pathogenesis of various retinal diseases.

One type of biomolecule found in vitreous humor is microRNA (miRNA), a class of small non-coding RNA that regulates gene expression. For example, miRNAs may silence gene transcription under certain conditions by hybridizing target messenger RNA (mRNA) to prevent protein translation or tag the mRNA for degradation (16). Extracellularly, miRNAs mediate communication between cells, with effects ranging from development of mesenchymal stem cells (17), to apoptosis of vascular cells (18). A benefit of miRNAs as an extracellular entity, in contrast to other forms of RNA or proteins, is its size. Averaging 21-25 nucleotides, miRNAs are small enough to avoid nucleases and resist changes in heat and pH across different bodily fluids (19). This characteristic has allowed miRNAs to serve as a potential biomarker for many hematologic and solid cancers (20), temporal lobe epilepsy (21), acute myocardial infarction (22), and sepsis (23).

Of the extracellular miRNAs found in vitreous humor, approximately 10% are secreted through exosomes (24). Exosomes are extracellular vesicles, typically 40-160 nm, derived from endosomes and secreted by most cell types, including those of the retina (25). Exosome vesicles contain cell constituents such as proteins, lipids, RNA, and DNA, and can release their contents into neighboring or distant target cells (26). Although the exact function of exosomes is unknown, this targeted transfer of contents suggests that they are involved in intercellular communication. The potential roles of exosomes span widely across wound healing, immunity, cellular differentiation, and more (25). Exosomes have been isolated in most bodily fluids, including blood, saliva, urine, and cerebrospinal fluid, and the contents of these exosomes have been found to be altered by disease (27). For example, serum levels of miR-21 have been significantly increased in patients with esophageal and hepatocellular carcinoma (28, 29). Similarly, salivary exosome levels of miR-1246 and miR-4644 are significantly higher in pancreato-biliary tract cancers, and the combined levels of these miRNAs have potential as a sensitive screening tool for these cancers (30). Furthermore, exosomal miR-142-5p and miR-223 were found to be significantly upregulated in breast milk as an early detector of intramammary infection (31). However, while serum, saliva and breast milk are accessible through safe and relatively simple collection methods, other fluids like cerebrospinal fluid and vitreous require more invasive techniques and more advanced clinical skills, therefore limiting their accessibility for research, and almost completely precluding the possibility of collecting control samples from healthy counterparts.

In this meta-analysis, we searched the current literature for peer-reviewed scientific evidence linking changes in vitreous composition with retinal disease, focusing on miRNA characterization and quantification. Out of 126 studies that were screened, we found 22 that specifically addressed the question of miRNAs in vitreous biopsies of retinal disease patients, and reanalyzed the data they provide. Overall, our meta-analysis shows that vitreal miRNAs do serve as useful biomarkers in certain retinal diseases such as proliferative diabetic retinopathy, age-related macular degeneration, and proliferative vitreoretinal disease. However, the consistency of data regarding miRNA content in human vitreous is limited by the fact that no gold standard for miRNA purification and amplification has yet been established. Additionally, studies lack a source of true control samples as vitreous can only be collected when a patient is undergoing vitrectomy, and patients only undergo vitrectomy when their eye is in some state of disease. In recognition of these challenges, we propose using certain housekeeping genes and data normalization techniques to standardize the analysis of miRNA in the vitreous, and we explore potential methodologies for obtaining vitreous samples from healthy patients. Finally, we identify miR-142, miR-9, and miR-21 as the strongest candidates for use as biomarkers of certain reginal diseases. Specifically, these miRNAs were upregulated in proliferative diabetic retinopathy and proliferative vitreoretinal disease.

## Main Text

### 1. Meta-analysis methodology

#### Search methods for identifying studies

Studies were identified via PubMed search with the search terms “exosome vitreous” and “microRNA vitreous” (**Figure 1**). The scope of articles was limited to studies published after 2012, considering that there were no studies describing miRNA in vitreous humor prior to 2013 (12). The last search was performed on January 23, 2024. All studies were reviewed by one reviewer, and selected studies were confirmed by a second reviewer with subsequent resolution of conflicting ideas.

**Figure 1.**
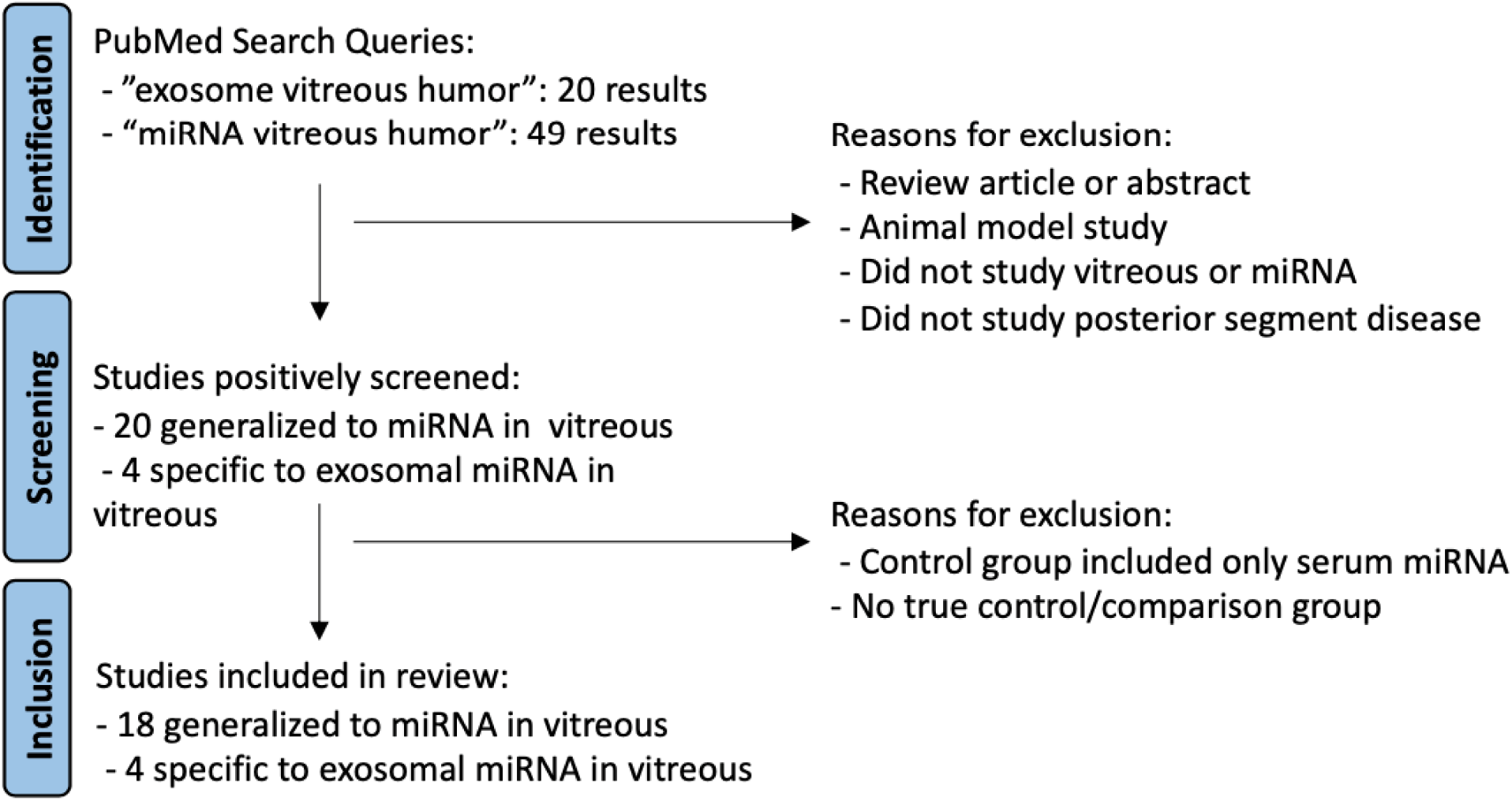
Flowchart describing search methodology and exclusion criteria.

#### Eligibility criteria

All selected studies were peer-reviewed, primary research articles reporting original data. Only studies that collected vitreous samples from human subjects were included. Selected studies researched diseases that directly affect the retina, including primary retinal pathologies and metastases to the retina, but excluded anterior segment diseases. With regards to methodology, all selected studies performed miRNA isolation using a microarray kit from a tested manufacturer, such as Qiagen, Exiquon, or TRIzol, followed by miRNA quantification with either qPCR, next generation sequencing (NGS), or a microarray scanner. All studies must have tested significance of differential expression of miRNA, either exosomal or non-exosomal, against control samples that were also derived from vitreous humor. Studies that had a sample size less than 2 for any cohort were excluded. Studies that did not show evidence of miRNAs in the vitreous were excluded as well.

#### Data synthesis and analysis

Exosomal and non-exosomal miRNAs that were found to be dysregulated in disease were identified within the selected studies as well as in any supplementary tables. Gene Ontology (GO) and Kyoto Encyclopedia of Genes and Genomes (KEGG) analysis were conducted on these miRNAs using the DIANA mirPath v.4 pathway analysis web server, analyzing the most commonly dysregulated miRNAs for disease categories that had at least five relevant studies (32). When searching for commonly dysregulated miRNAs, we first studied individual isoforms and then grouped them by mature miRNA family. Statistical analysis was conducted through the DIANA mirPath v.4 software, using the “classical analysis” option, which uses one-sided Fisher’s Exact tests to determine significance (p<0.05).

### 2. Meta-analysis results

#### Vitreous humor sampling and molecular isolation

The studies assessing the potential of vitreous biopsies as a diagnostic tool included various diseases but were limited to populations requiring vitrectomies. The studies also varied significantly in their miRNA extraction and purification techniques due to the lack of a gold standard among the many commercially available technologies designed to isolate and purify specific types of RNA.

The vitreous humor is routinely extracted in humans through an invasive procedure known as a vitrectomy. Vitrectomies are routinely performed as treatment for RD, macular holes (MH), ERM, posterior vitreous detachment, and vitreous hemorrhage. Vitrectomies may also be necessitated due to complications in cataract surgery, such as posterior capsule rupture, which occurs in 0.45-7.9% of cataract surgeries (33), and in patients with dislocated intraocular lens (34). While vitrectomies must be performed in the operating room, vitreous humor can also be safely sampled in the clinic prior to intravitreal injections, which are commonly done for the treatment of diabetic retinopathy, neovascular AMD, and retinal vein occlusions (35). However, in-office sampling is not routine, and the studies reviewed were thus limited to subjects who required vitrectomies as treatment. As such, control subjects did not have non-pathological eyes but rather required a vitrectomy for visual disturbance from conditions such as idiopathic floaters, MH, ERM. Therefore, control samples were defined as obtained from patients with minimal risk of visual loss progression and blindness, as compared to the studied diseases. The invasiveness of the vitrectomy limited the sample sizes of all studies.

After collecting the vitreous humor, all the studies, except those done by Liu et al. and Tuo et al., specified that the vitrectomy samples were immediately stored at -80 °C prior to experimentation, to prevent RNA degradation. After the frozen vitreous samples were thawed, miRNAs were isolated using commercially available purification kits. The volume of vitreous humor collected through vitrectomy, varies from study to study, and within patients in the same study, but was typically 1-5 ml. Although there are no RNA isolation kits specifically designed for vitreous humor, studies have compared the efficacy of various kits for use in small samples of other body fluids such as plasma and urine. For example, TRIzol reagent (Invitrogen) has lower yield for miRNA with low guanine and cytosine content (36). However, the results of TRIzol extraction aligns most closely with miRNA documented in the miRBase database compared to miRNeasy mini kit (Qiagen) (37). Cell count and density of the sample also impact which isolation kit is preferred. For miRNA extracted from cells in culture, both miRNeasy and TRIzol had greater RNA yields than miRCURY (Qiagen) at lower cell counts, and miRNeasy outperformed both TRIzol and miRCURY at both low and high cell densities (38). While a gold standard for miRNA isolation in vitreous humor has yet to be established, the variability across isolation methods in other biofluids suggests a similar potential for variability in vitreous humor, which may account for some of the differences across the reviewed studies. Of the reviewed studies that isolated total miRNA, 13 used miRNeasy, 4 used TRIzol and 1 used miRCURY.

To extract exosomal miRNAs, an additional step is required to isolate the exosomes first. Exosomes can either be isolated from vitreous humor via a combination of ultracentrifugation and filtration, known as differential centrifugation, or via centrifugation followed by an exosome isolation kit such as miRCURY Exosome Kit (Qiagen) or RiboTM Exosome Isolation Reagent (RiboBio). Differential centrifugation is considered the gold standard for exosome isolation, and isolation kits may lead to the inclusion of non-exosomal particles (39). However, ultracentrifugation requires costly equipment that isolation kits avoid. Of the reviewed studies which isolated miRNA from exosomes, two used differential centrifugation, while two employed an isolation kit.

#### miRNA changes in total vitreous biopsy across diseases

The reviewed studies varied in their level of specificity when reporting dysregulated miRNA expression. For example, some reported the mature miRNA family that was dysregulated (e.g.: miR-125) while others also included a suffix (i.e.: -3p, -5p), specifying an isoform within the miRNA family. miRNA isoforms were considered separate molecules and are denoted by different letters (e.g.: miR-125a vs miR-125b). When searching for the most commonly dysregulated miRNAs across all of the studies, we included both the overall miRNA families, as well as individual isoforms.

With regards to the miRNA families, miR-16 was most commonly dysregulated, with 9 occurrences in 7 different studies (**Table 1**). miR-16 was upregulated in all instances except in the case of PM, where it was downregulated. This may reflect the pathogenesis of PM compared to the other diseases. While PM is characterized by chorio-retinal atrophy, all the other pathologies involved proliferative or cancerous processes. In addition, the study on PM included controls that had RRD. The inflammatory and immune reaction to RRD could cause miRNA dysregulation, making miR-16 relatively downregulated in subjects with PM. Other commonly dysregulated miRNA families include miR-125b, miR-139, and miR-423, as described in **Table 1**.

**Table 1.**
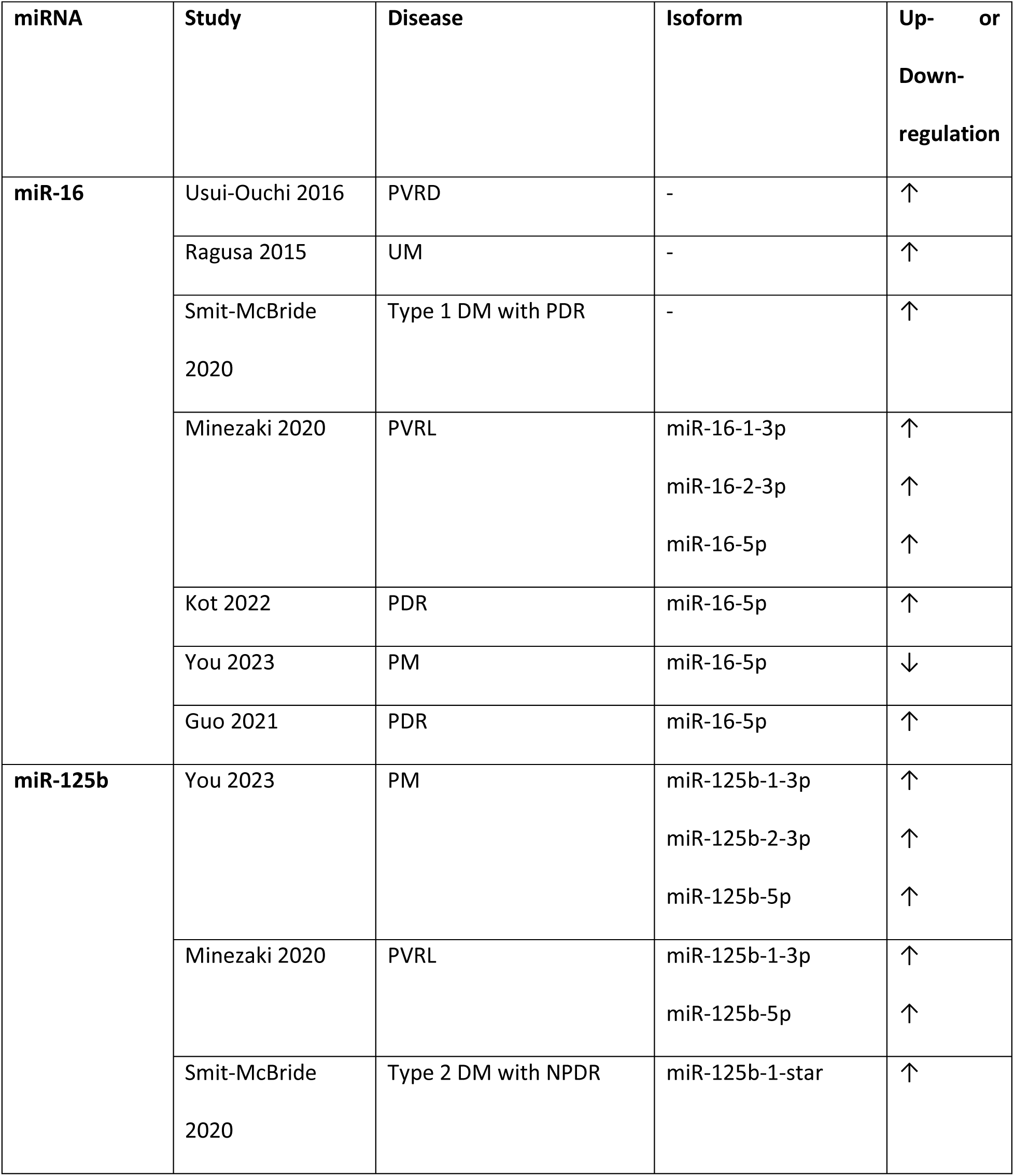
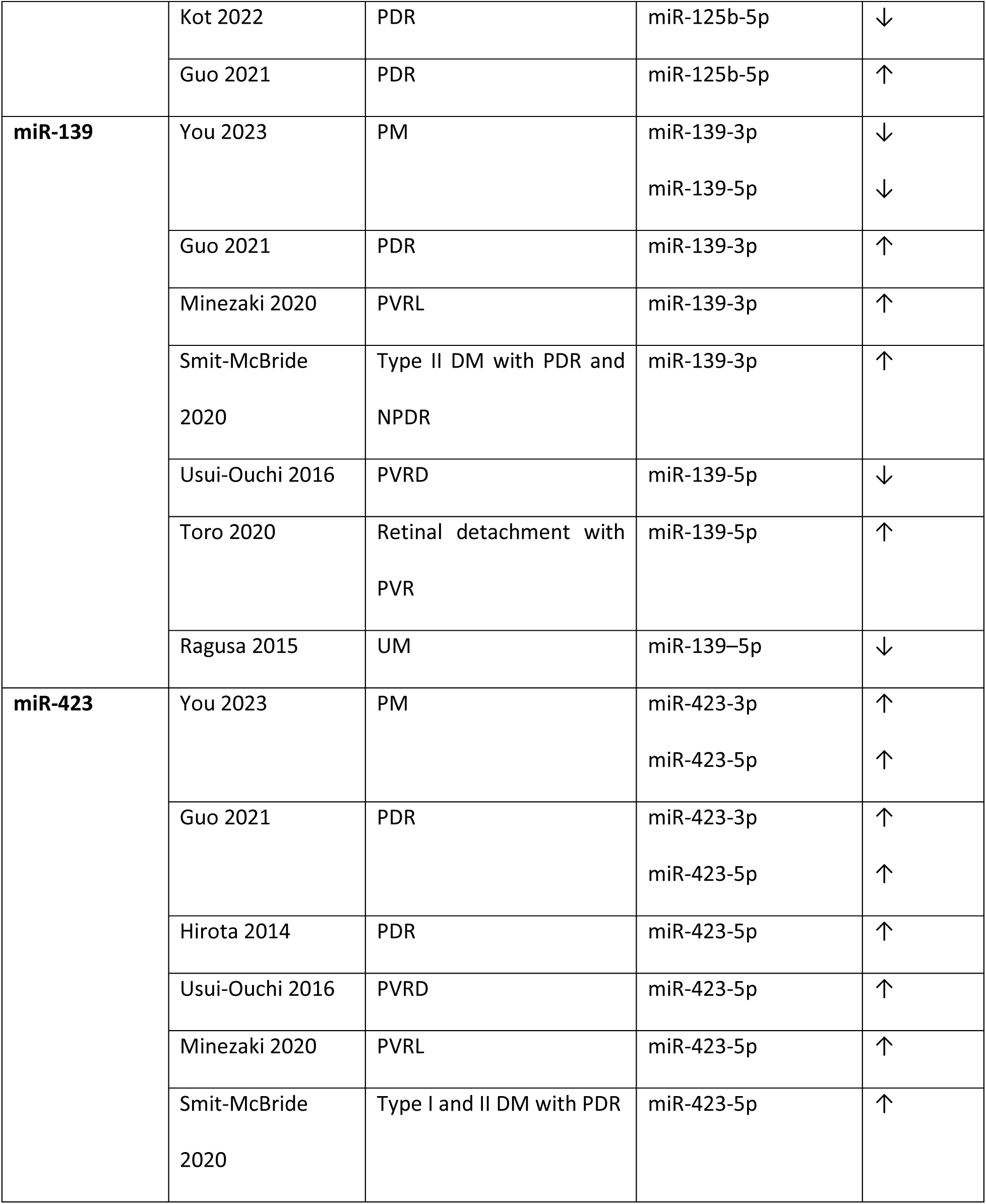
Correlation between changes in human vitreous miRNA and retinal diseases. The table shows the most commonly dysregulated miRNA across all diseases included in the present meta-analysis. PVRD: proliferative vitreoretinal disease, UM: uveal melanoma, DM: diabetes mellitus, PDR: proliferative diabetic retinopathy, PVRL: primary vitreoretinal lymphoma, PM: pathologic myopia, NPDR: non-proliferative diabetic retinopathy, PVR: proliferative vitreoretinopathy

Comparing specific isoforms, miR-423-5p was most frequently dysregulated with 6 different studies finding it to be upregulated, as seen in **Table 1**. The next most frequently dysregulated miRNA was miR-320b, which does not have -3p or -5p isoforms according to the miRBase database. miR-320b was consistently upregulated in 5 different studies investigating PDR, PM, and primary vitreoretinal lymphoma (PVRL) (40–44).

#### miRNA in diabetic retinopathy

DR can be categorized as NPDR, the less severe form, or PDR, the more advanced form, which is characterized by breakdown of the barrier between the retina and its surrounding blood vessels (3). Of the ten articles in our analysis that studied DR, only one studied NPDR while the remaining nine focused on PDR. Pramanik et al. compared miRNA in Type 2 DM patients with and without NPDR and found that NPDR patients had significantly decreased levels of miR-126 and miR-132 in both the vitreous humor and plasma (45). Furthermore, ROC curve analysis showed that the fold change of miR-126 and miR-132 in the vitreous humor had greater sensitivity and specificity as a predictor of NPDR compared to the fold change in plasma, indicating that the vitreous miRNAs have better diagnostic utility for retinal disease than serum miRNA (45). Of the seven studies in our analysis that researched PDR, the two studies by Liu et al. and Kot et al. specifically isolated exosomal miRNA and are detailed further in section 3.3.1 below, while the remaining seven articles studied miRNA found in the total vitreous humor.

Given that abnormal blood vessel growth is part of the pathogenesis of PDR, several of the studies correlated miRNA that were dysregulated in PDR with angiogenesis. For example, Hirota et al. identified five miRNAs (miR-15a, miR-320a, miR-320b, miR-93, and miR-29a) that were significantly elevated in the vitreous humor in patients with PDR and thought to be involved in angiogenesis (40). For example, miR-15a has been shown to inhibit angiogenesis in response to ischemia by reducing the activity of fibroblast growth factor 2 and VEGF, indicating that miR-15a may have protective effects in PDR (46). Guo et al. identified three miRNAs (miR-24-3p, miR-197-3p, and miR-3184-3p) that were not only significantly upregulated in PDR but also correlated with increased levels of VEGF and transforming growth factor-β (TGFβ) in the vitreous humor (43). In contrast, Gomaa et al. showed that miR-200b was elevated in PDR but it did not correlate with vitreous levels of VEGF (47). Yang et al. identified seven miRNA that were elevated in PDR, and KEGG analysis showed that the target genes of these miRNA were most enriched in the Th17 cell differentiation pathway (48). Notably, interleukin-17, which is produced by Th17 cells, has been shown to promote retinal neovascularization through multiple mechanisms (49–51).

The high glucose conditions caused by diabetes also contribute to the pathogenesis of PDR, Shanbagh et al. specifically studied how these conditions affected miRNA and target protein levels (52). They found that miR-182-5p was upregulated in patients with PDR, and under high glucose conditions, RPE cells had higher expression of miR-182-5p. The study also showed that the downstream effects of elevated miR-182-5p expression included downregulation of FoxO1 and upregulation of Akt and HK2, which are known to be involved in metabolic regulation (53, 54). These findings show that miR-182-5p may have a role in the pathogenesis of PDR through its effects on glucose metabolism.

Smit-McBride et al. differentiated between Type 1 and Type 2 DM when studying miRNA in the vitreous of patients with PDR. They identified four miRNA families (let-7, miR-320b, miR-4488, and miR-762) that were upregulated in patients with PDR with both types of DM as compared to controls with MH or macular pucker (41). Although these miRNAs have not been studied in the retina, let-7 targets regulatory cytokines that promote angiogenesis and has been linked to increased risk of Type 2 DM (55, 56). Interestingly, miR-320b has been shown to be anti-angiogenic via its regulation of neuropilin 1 and insulin-like growth factor 1, indicating that the increased levels of miR-320b in PDR may be a compensatory mechanism attempting to attenuate disease progression (57, 58).

Because the retinal blood vessels that form in PDR are abnormal, they are at high risk of bleeding into the neighboring vitreous humor. Mammadzada et al. were the only researchers to specifically look at PDR that was complicated by recurrent vitreous hemorrhage. They found that miR-20a and miR-93 were significantly elevated in patients who later developed recurrent vitreous hemorrhage, and both miRNA are known to be involved in angiogenesis. Increased levels of miR-20a have been shown to downregulate VEGF protein expression, but miR-20a is also known to be downregulated by hypoxic conditions (59). Of note, miR-20a was also found to be elevated in the study by Friedrich et al. (60). miR-93, which was also found to be upregulated in the study by Hirota et al., has been shown to modulate angiogenesis through multiple mechanisms, including by directly binding the VEGF-A gene and by promoting endothelial cell proliferation and migration by targeting cyclin-dependent kinase inhibitor 1A (59, 61, 62)

Given that VEGF is considered a primary driver of neovascularization in PDR, anti-VEGF medications have become a standard treatment and have been shown to improve visual acuity in PDR (63). Friedrich et al. not only identified miRNA that were elevated in PDR but also found that miR-23b-3p levels were subsequently lowered after anti-VEGF treatment (60). Further investigation into the targets of miR-23b-3p is needed. Overall, there were three families of mature miRNAs that were found to have four isoforms dysregulated in at least three of the nine studies, as shown in **Table 2**.

**Table 2.**
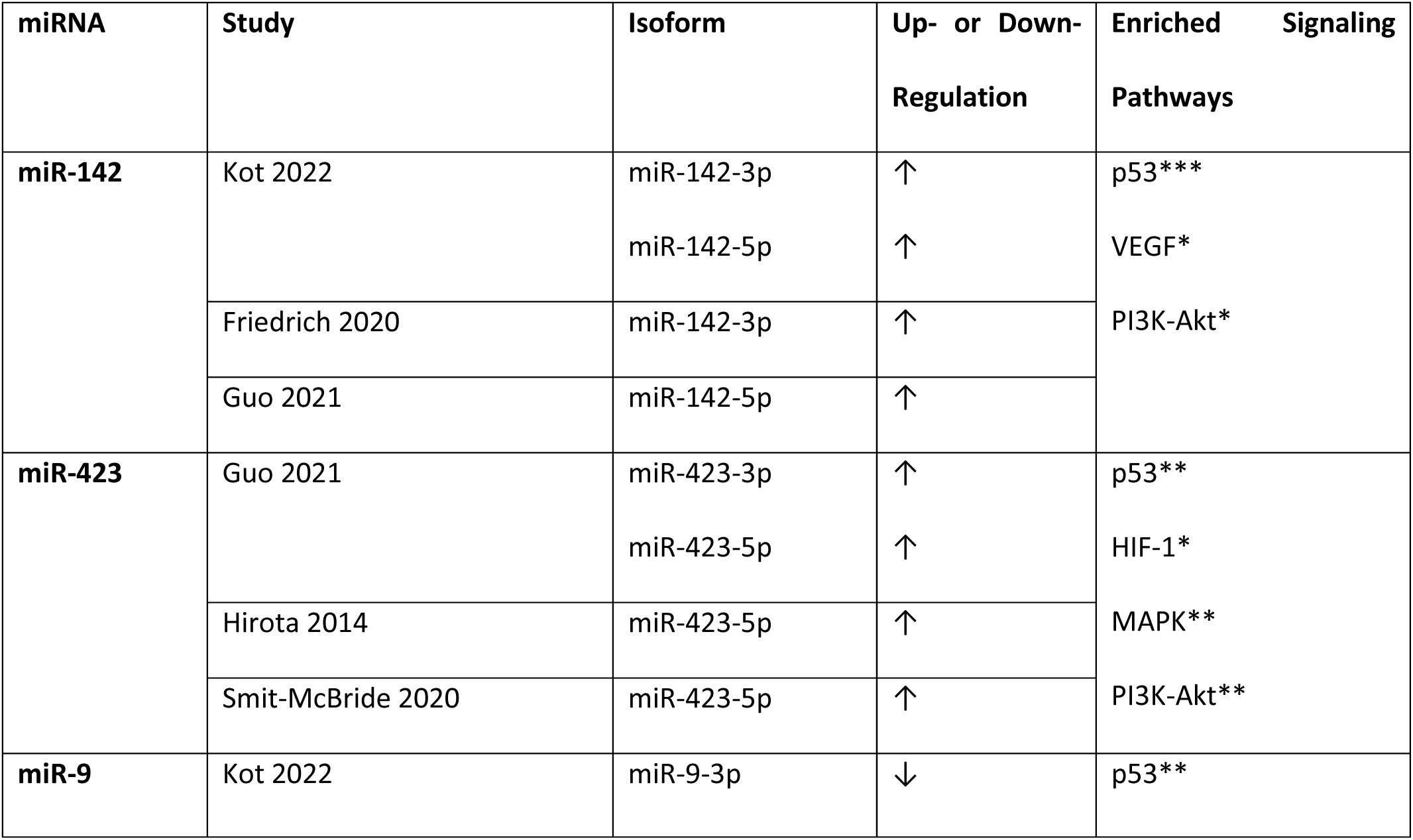
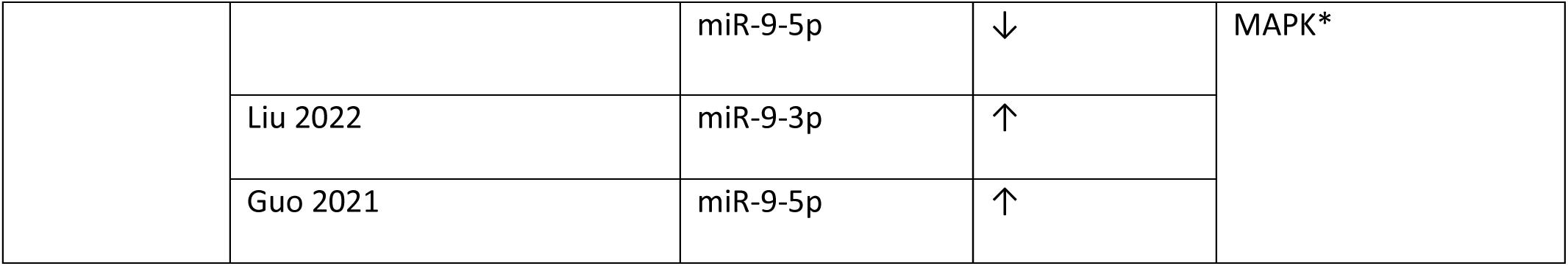
Correlation between changes in human vitreous miRNA expression and PDR. The table shows the miRNA families with °4 instances of dysregulation, across studies, in PDR; and the molecular signaling pathways they affect related to diabetes, with or without retinopathy. *p<0.01, **p<0.001, ***p<0.0001.

KEGG analysis through the DIANA-miRPath database showed that all three miRNA families (miR-142, miR-423, and miR-9) were connected to pathways in diabetes and diabetic retinopathy. For example, all three are involved in the p53 signaling pathway, which is known to be connected to Type 2 Diabetes Mellitus (DM), because p53 inhibition preserves pancreatic insulin secretion (64). Furthermore, p53 protein and mRNA have been shown to be upregulated in retinal pericytes and human retinal vascular endothelial cells under high glucose conditions, showing a possible role in the early development of diabetic retinopathy (65, 66). One limitation of KEGG analysis is that although it can determine whether a miRNA targets a certain pathway, it does not specify whether the pathway is enhanced or suppressed by the miRNA. In this case, if the miRNA were shown to upregulate the p53 signaling pathway, that would provide further evidence in their role in the pathogenesis of PDR.

The VEGF signaling pathway is considered a primary driver for the vascular proliferation that characterizes PDR (67, 68). Only miR-142 directly enriched the VEGF signaling pathway, but both miR-142 and miR-423 enriched related pathways. For example, miR-423 targeted the hypoxia-inducible factor (HIF)-1 signaling pathway, which is known to increase retinal VEGF in diabetic retinopathy. As its name suggests, HIF-1 is induced by small vessel hypoxia in the retina created by diabetes and then mediates retinal neovascularization through the release of VEGF and erythropoietin (69–72). Therefore, miRNA that activate HIF-1 signaling would theoretically worsen diabetic retinopathy while miRNA that suppress it could be protective. Further research is required to determine how miR-423 levels correlate with HIF-1. Similarly, miR-142 and miR-423 targeted the phosphatidylinositol 3-kinase (PI3K)-Akt signaling pathway. Activation of PI3K-Akt can increase VEGF expression by upregulating the HIF-1 signaling pathway or by phosphorylating endothelial nitric oxide synthase, which is a necessary step in VEGF-induced angiogenesis (73, 74). miRNA activation of the PI3K-Akt pathway would increase neovascularization in PDR. Both miR-423 and miR-9 enriched the MAPK signaling pathway, which is activated by the VEGF signaling pathway, and is known to cause retinal inflammation and fibrosis in PDR (75). In fact, MAPK inhibition in retinal mouse models decreased inflammatory and angiogenic markers such as nitric oxide and cyclooxygenase-2 (76, 77). Upregulation of MAPK by miRNA would thus contribute to the development of PDR.

When evaluating the use of the miRNAs as biomarkers for PDR, both their specificity and consistency must be considered. Although miR-423 is consistently found to be upregulated in PDR, it is also commonly upregulated in other disorders such as PM and PVRL (**Table 1**). This lack of specificity for PDR indicates that miR-423 may be a more general marker of retinal cell dysfunction. On the other hand, although miR-9 was not found to be dysregulated as frequently as miR-423, its results were inconsistent. Forms of the miR-9 family were downregulated in the study by Kot et al. but upregulated in the studies by Liu et al. and Guo et al (43, 78, 79). These differences may be due to variations in subject choice and study design. In the study by Kot et al., the PDR subjects had disease that was complicated by tractional RD, meaning the PDR was severe enough to produce a fibrous proliferative membrane that created enough traction to cause detachment of the neurosensory layer of the retina (80). In the other two studies, the PDR was not complicated by tractional RD, and the RD and its treatment may have impacted the miRNA composition. Also, Kot et al. used qPCR to quantify assayed miRNA while Liu et al. and Guo et al. used NGS. Although Guo et al. verified the results of their NGS with qPCR, the initial choice of miRNA quantification technique may have led to varied results. Until these discrepancies can be resolved, the utility of miR-9 as a biomarker is limited. miR-142, however, was consistently upregulated across studies and was more specifically upregulated in the setting of PDR. This, combined with its direct and indirect influence on the VEGF pathway, makes miR-142 an important potential biomarker for PDR.

The three most commonly dysregulated miRNA families, with at least four instances of dysregulation in at least three different studies, listed in **Table 2** as well as the 16 next most commonly dysregulated miRNA families, with at least three instances of dysregulation (Supp. Table 1) were selected for pooled GO and KEGG analysis (see **Figure 2 & 3**). KEGG analysis resulted in 141 related pathways. Notably, ubiquitin-mediated proteolysis, protein processing in the endoplasmic reticulum, cell cycle, and p53 signaling pathways, are all related to Type 2 DM according to the KEGG pathway database. GO analysis revealed 1174 targeted categories, with 547 results in the biological component domain, 163 in molecular function, and 245 in the cellular component.

**Figure 2.**
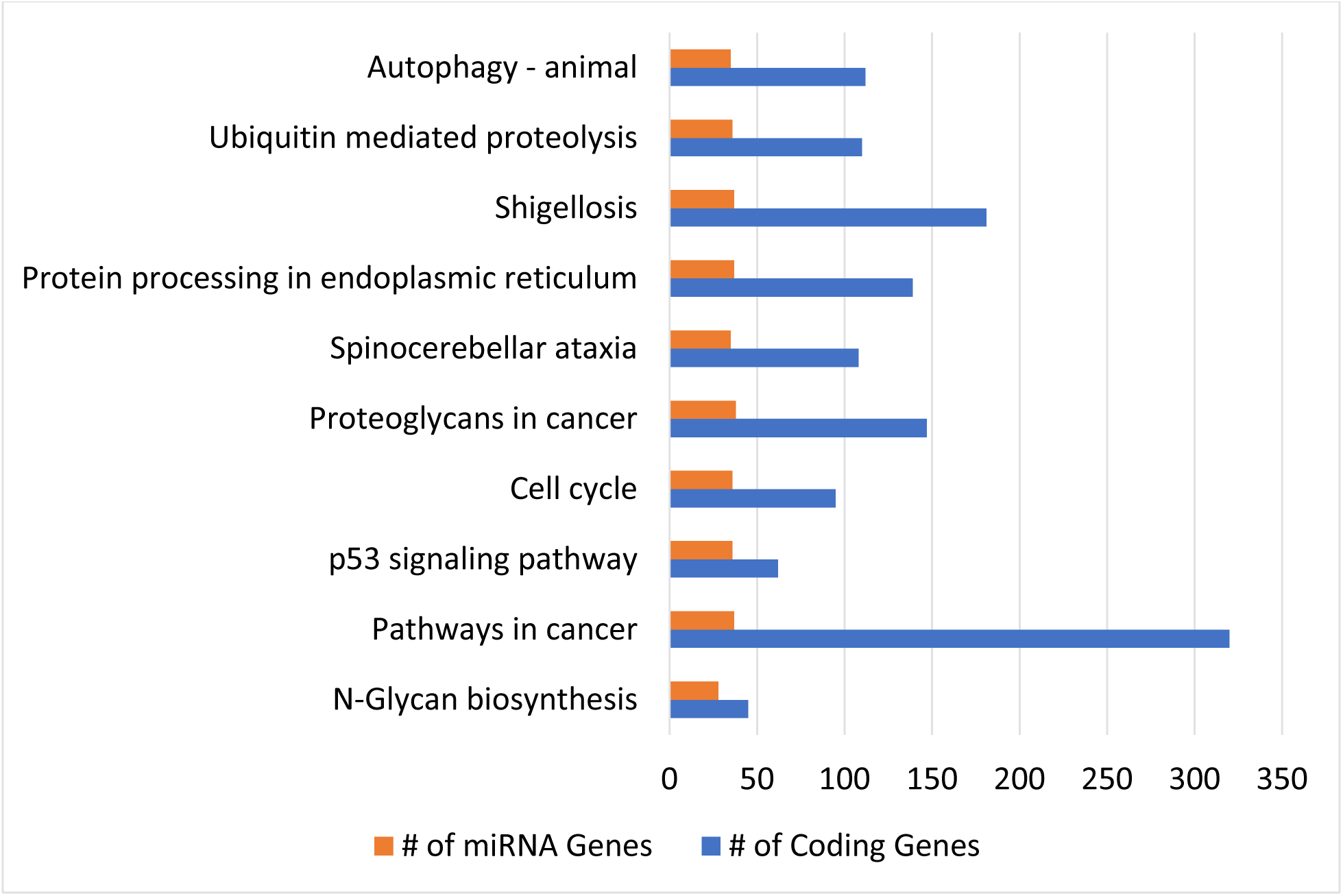
KEGG analysis of human vitreous miRNAs in PDR. Top 10 most enriched KEGG pathways for the 19 most commonly dysregulated (°3 instances of dysregulation) miRNA species in proliferative diabetic retinopathy, including 41 isoforms. The miRNA number indicates the number of inputted miRNAs that are involved in each pathway and the gene number indicates the number of experimentally validated target coding-genes of the inputted miRNAs that are involved in each pathway. The top 10 pathways had p-values <1E-11 (one-sided Fisher’s exact test, p<0.05).

**Figure 3.**
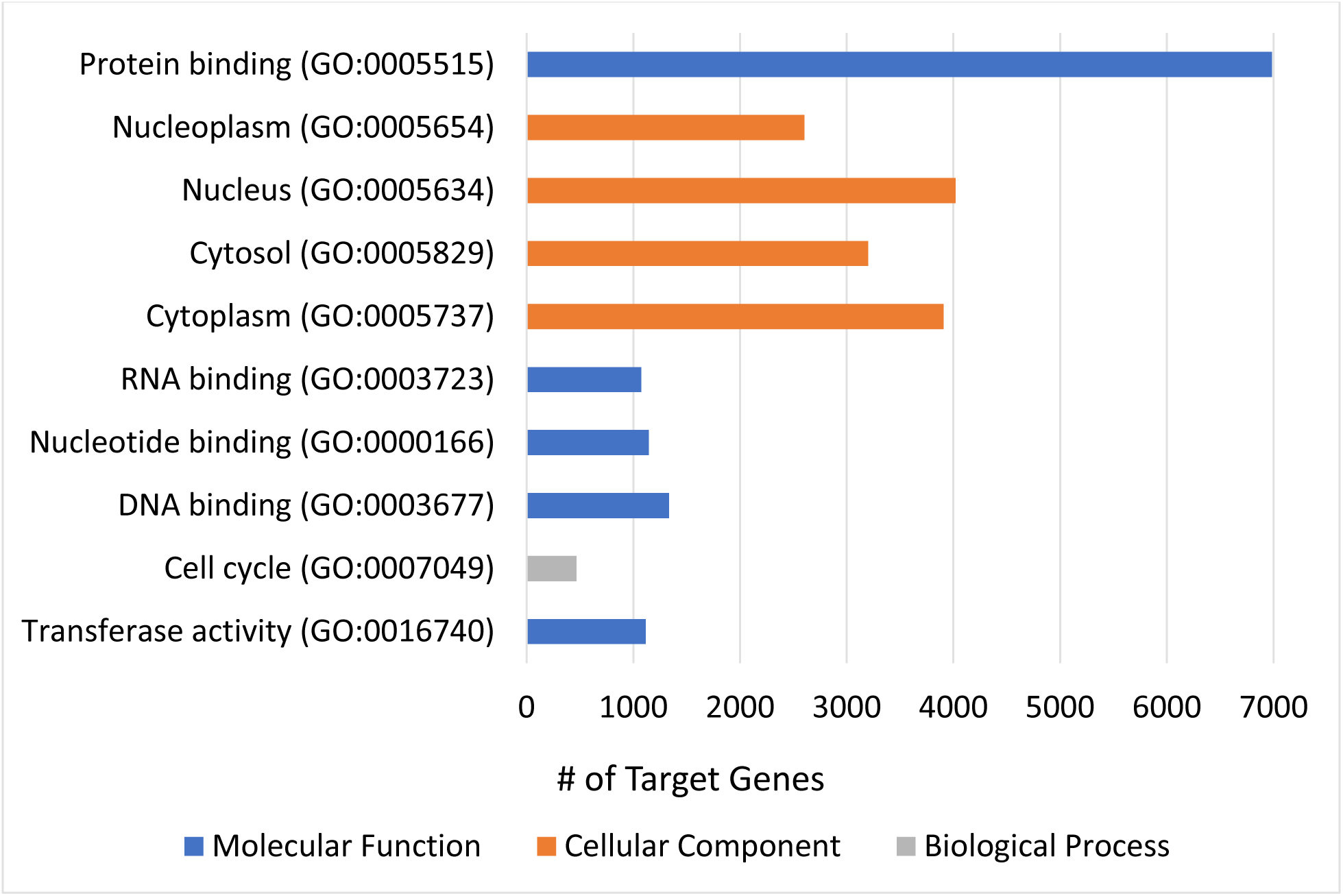
GO-terms analysis of human vitreous miRNAs in PDR. Top 10 most enriched GO terms for the 19 most commonly dysregulated (°3 instances of dysregulation) miRNA species in proliferative diabetic retinopathy, including 41 isoforms. The top 10 terms had p-values <1E-49 (one-sided Fisher’s exact test, p<0.05).

#### miRNA in proliferative vitreoretinal disease

Proliferative vitreoretinal disease (PVRD) is a category encompassing diseases with a proliferative element to their pathogenesis, including PVR following trauma or RD and PDR. In our analysis, the study by Usui-Ouchi et al. encompassed both PDR and PVR, and Toro et al. focused on PVR due to RD (81, 82). We identified 25 of the most commonly dysregulated miRNAs (with at least 3 instances of dysregulation) among these two studies as well as the 9 looking at PDR alone (Supp. Table 2). As with PDR alone, the miR-9 and mir-423 families were two of the most commonly dysregulated, with miR-9-5p upregulated in PVR and miR-423-5p upregulated in general PVRD. GO and KEGG analysis of these 50 miRNAs showed almost complete overlap with the results from PDR alone. However, one notable difference was that the p53 signaling pathway was not in the top 10 most enriched pathways for PVRD although it was for PDR alone. As noted above, the p53 signaling pathway is known to be related to the pathogenesis of Type 2 DM, so it is reasonable that this pathway would be less targeted in PVR, a non-diabetic process. Instead, the neurotrophin-signaling pathway was in the top KEGG pathways for PVRD, although it was not among the most enriched pathways in PDR.

Usui-Ouchi et al. argue that the shared proliferative process in PDR and PVR would lead to similar alterations in miRNAs. In particular, they highlight miR-21 as increasing in relation to retinal fibrosis in PVRD. In fact, miR-21-5p is found to be upregulated in the studies by Guo et al. and Kot et al looking at PDR. However, miR-21-3p is downregulated in the PVR study by Toro et al. This discrepancy highlights the need for specification of miRNA isoforms across studies. Although both isoforms have the same mature miRNA, the differences in their precursors could affect how they correlate to pathology. In the past, studies may have been limited by the available microarrays, but moving forward, researchers should aim to specify the miRNA isoforms.

Another miRNA that may be involved in the pathogenesis of PVR is miR-148a-3p, which was found to elevated in patients with retinal detachments (83). Although the study subjects did not yet have progression to PVR, Takayama et al. showed that transfection of miR-148a-3p into human retinal pigment epithelium (RPE) cells promoted epithelial–mesenchymal transition, a process that characterizes PVR (83, 84).

#### miRNA in primary vitreoretinal lymphoma

Two studies, by Tuo et al. and Minezaki et al., identified miRNA that are dysregulated in primary vitreoretinal lymphoma (PVRL) in comparison to uveitis (44, 85). Because PVRL can be mistaken for uveitis, leading to delay in proper treatment, there is need for a distinguishing biomarker. The miRNAs found to be dysregulated in PVRL compared to uveitis in both studies can be found in **Table 3**, but only the results from Minezaki et al. are statistically significant.

**Table 3.** Dysregulated human vitreous miRNA in PVRL, as compared to uveitis. Only the results reported by Minezaki et al. were statistically significant (t-test, *p<0.05).

| miRNA | Study | Tail | Up- or Downregulation |
| --- | --- | --- | --- |
| **miR-200c** | Tuo 2013 | - | ↓ |
|  | Minezaki 2020 | miR-200c-5p | ↑* |
| **miR-484** | Tuo 2013 | - | ↑ |
|  | Minezaki 2020 | - | ↑* |
| **miR-197** | Tuo 2013 | - | ↑ |
|  | Minezaki 2020 | miR-197-3p | ↑* |
|  | Minezaki 2020 | miR-195-5p | ↑* |
| **miR-132** | Tuo 2013 | - | ↑ |
|  | Minezaki 2020 | miR-132-3p | ↑* |

Tuo et al. focused on miR-155, which they not only found to be significantly decreased in lymphoma patients but was also inversely correlated with IL-10, which is known to be secreted heavily in B-cell PVRL. However, miR-155 was not found to be dysregulated by Minezaki et al. One reason for the variation in significance and miRNAs identified between the two studies may be their sample sizes. Tuo et al. had only 3 PVRL and 3 uveitis samples for the microarray while Minezaki et al. had 6 each. However, Tuo et al. confirmed the results of the microarray with individual assays and qPCR from a larger sample of 17 PVRL and 12 uveitis subjects. Minezaki et al. used a microarray scanner to quantify the miRNAs and did not validate the results with qPCR. Also, the choice of microarray may have impacted the comprehensiveness of study results. The microarray used by Tuo et al. (the microRNA-CURY panel from Exiquon) tested for only 168 miRNAs while the 3D-Gene miRNA Oligo Chip used by Minezaki et al. tested for 2565 miRNAs. Further studies comparing miRNA in PVRL and uveitis should employ large-scale microarrays as done by Minezaki et al. but with the secondary validation schema outlined by Tuo et al.

#### miRNA in pathologic myopia

PM is defined by an axial length of at least 26.5 mm or a refractive error of at least -6.00 diopters. Patients with PM are at risk for progressive vision loss due to chorioretinal atrophy as well as RD. Two studies in our review analyzed changes to non-exosomal and exosomal miRNA in the vitreous humor of subjects with PM. Ando et al. found that let-7c was upregulated in PM while miR-200a was downregulated (86). These results are confirmed by You et al., who found let-7c-5p to be upregulated while miR-200a-3p was downregulated in PM (42). Ando et al. argued that let-7c may be secreted by Müller glial cells migrating to the inner limiting membrane of the retina in highly myopic eyes. KEGG pathway analysis of miR-200a completed by Ando et al. showed that the let-7c was related to the PI3K/Akt signaling pathway, which may increase axial length by stimulating proteins such as insulin-like growth factor-1 and matrix metalloproteinase-2, which are known to be related to PM. You et al. identified 143-3p and miR-145-5p as key miRNA through weighted gene co-expression network analysis. Like Ando et al., You et al. showed that miR-143-3p and miR-145-5p may relate to axial length by potentially increasing the expression of insulin-like growth factor. One reason that Ando et al. may not have detected dysregulation of miR-143-3p and miR-145-5p is that these miRNAs were detected within exosomes. As discussed later in this review, exosomal miRNA can be more stable than non-exosomal miRNA and thus serve as more reliable biomarkers.

#### miRNAs in age-related macular degeneration

The pathogenesis of AMD involves several layers of the retina: the photoreceptor layer becomes less dense, there is hyperpigmentation and atrophy of the RPE, and lipid-rich materials known as drusen are deposited between the RPE and Bruch’s membrane (4). In the more advanced form, wet or neovascular AMD, there is neovascularization of the macula (4). Despite its prevalence, to the best of our knowledge, there is only one study looking at miRNA in the vitreous humor in neovascular AMD. Menard et al. noted upregulation of miR-146a and downregulation of miR-106b and miR-152 in patients with neovascular AMD compared to controls (87). The study goes on to argue that the ratio of miR-146a to miR-106b collected from both vitreous humor and plasma can be used as a biomarker for neovascular AMD, based on receiver operating characteristic (ROC) analysis. According to Menard et al., miR-146a may have a protective mechanism in the retina by downregulating inflammatory cytokines. Meanwhile, decreased miR-106b levels may worsen neovascular AMD by influencing VEGF-A and IL-8 regulation. This was the only study to consider the ratio of two miRNA as a biomarker, but while this approach is unique, it comes with limitations. miR-146a and miR-106b are proposed to have separate mechanisms for advancing neovascular AMD, so if one of these mechanisms progresses at a slower-than-expected rate, then the ratio will be skewed. This study’s sample size was 26, but a larger follow-up study could be used to verify the findings and establish a diagnostic range for the ratio.

#### miRNAs in intraocular tuberculosis

Intraocular tuberculosis (IOTB) can occur as a result of systemic tuberculosis or via direct infection of the eye. Although it most commonly presents as posterior uveitis, it can also affect the retina, usually through spread from the choroid, presenting as focal tubercles, subretinal abscesses, or diffuse retinitis (88). These findings are thought to be caused by an inflammatory response to bacteria-laden macrophages depositing their contents in the ocular blood supply (89). To our knowledge, only one study has analyzed vitreous-derived miRNA. Chadalawada et al. used NGS followed by validation with qPCR to identify 3 miRNA (miR-150-5p, miR-26b-5p, and miR-21-5p) that were upregulated in IOTB compared to controls with MH (90). Furthermore, the study used ROC analysis to identify a combination of miR-21-5p and miR-26b-5p as having the highest potential as a biomarker. Interestingly, the study noted that miR-21-5p may have immune regulated anti-inflammatory effects in IOTB, which contrasts with miR-21’s pro-fibrotic role in the pathogenesis in PVRD as discussed above. Given that miR-21 isoforms have been found to be dysregulated in 5 of our studies, it is important that further research is done to investigate its mechanism of action in retinal diseases.

#### Exosomes in disease

Exosomal miRNA may be even better markers of retinal disease than non-exosomal RNA. Exosomes are secreted into the vitreous humor by surrounding cells, such as retinal cells, providing miRNAs that reflect the state of the diseased parent cell (91). Exosomal miRNAs are protected from degradation by RNAses and show increased stability. In addition, exosomes have specific proteins and markers, and this uniqueness allows for more targeted analysis of low abundance exosomal cargo (91). Within the vitreous humor, exosomes are particularly good choice for finding biomarkers due to their abundance. Over 60% of proteins isolated from vitreous humor are consistent with exosomes, according to gene ontology analysis of vitreous humor proteins (92). Moreover, almost 50% of the vitreous humor proteins thought to be within the exosomes are associated with retinal disease, furthering the argument that exosomal cargo can be used as indicators of retinal health (92). In fact, studies have already shown that vitreous humor exosomes may affect immunomodulation and angiogenesis (93, 94). Here, we review how exosomal miRNA vary with disease and evaluate their potential as biomarkers.

#### Exosomal miRNA

Thus far, there are only four studies that have identified dysregulated miRNA in exosomes from the vitreous humor. The most notable miRNAs from each study are outlined in **Table 4**. Both Ragusa et al. and You et al. used differential ultracentrifugation, the gold standard, to isolate exosomes from vitreous humor, while Kot et al. and Liu et al. used a combination of centrifugation and an exosome isolation kit.

**Table 4.**
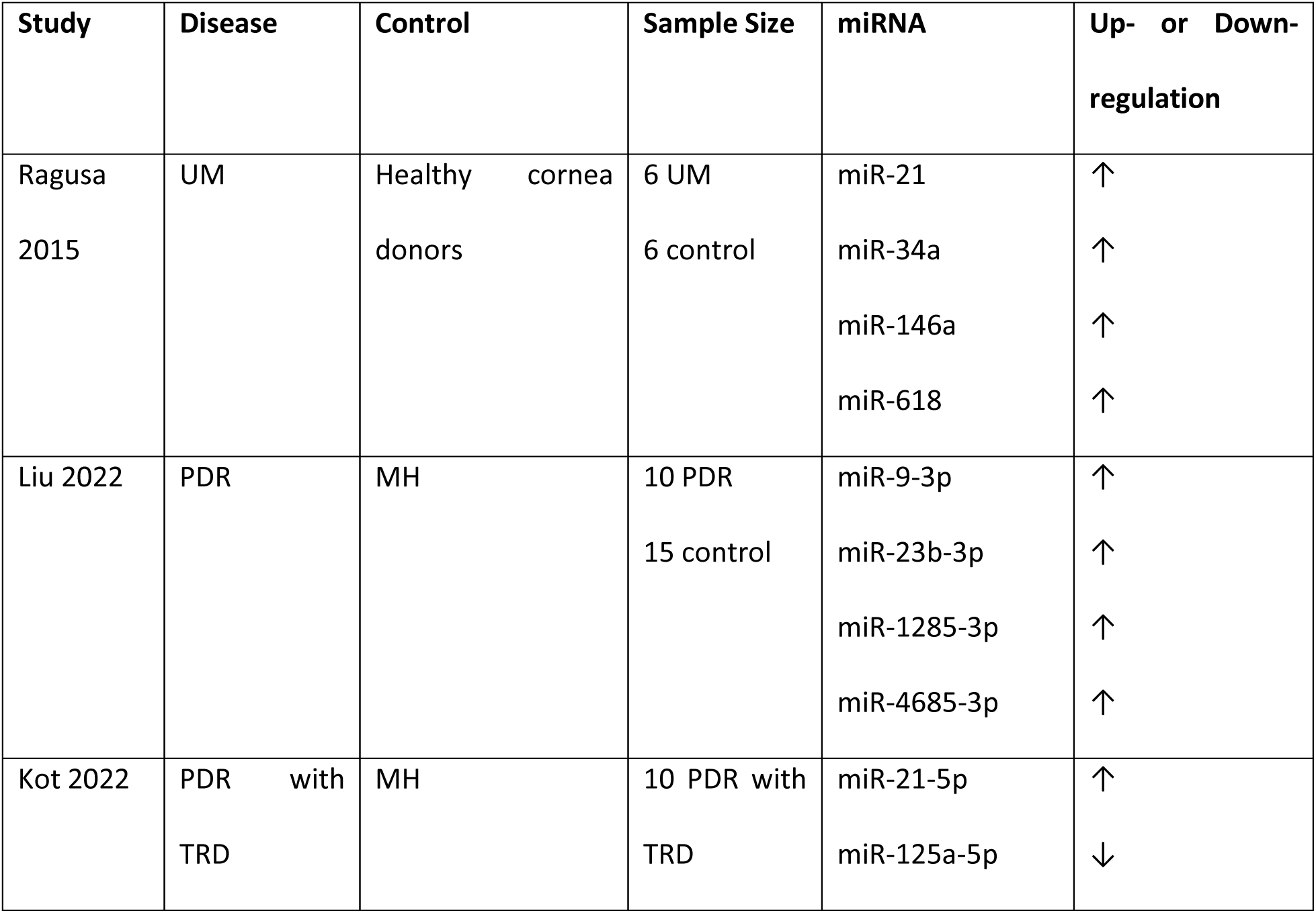
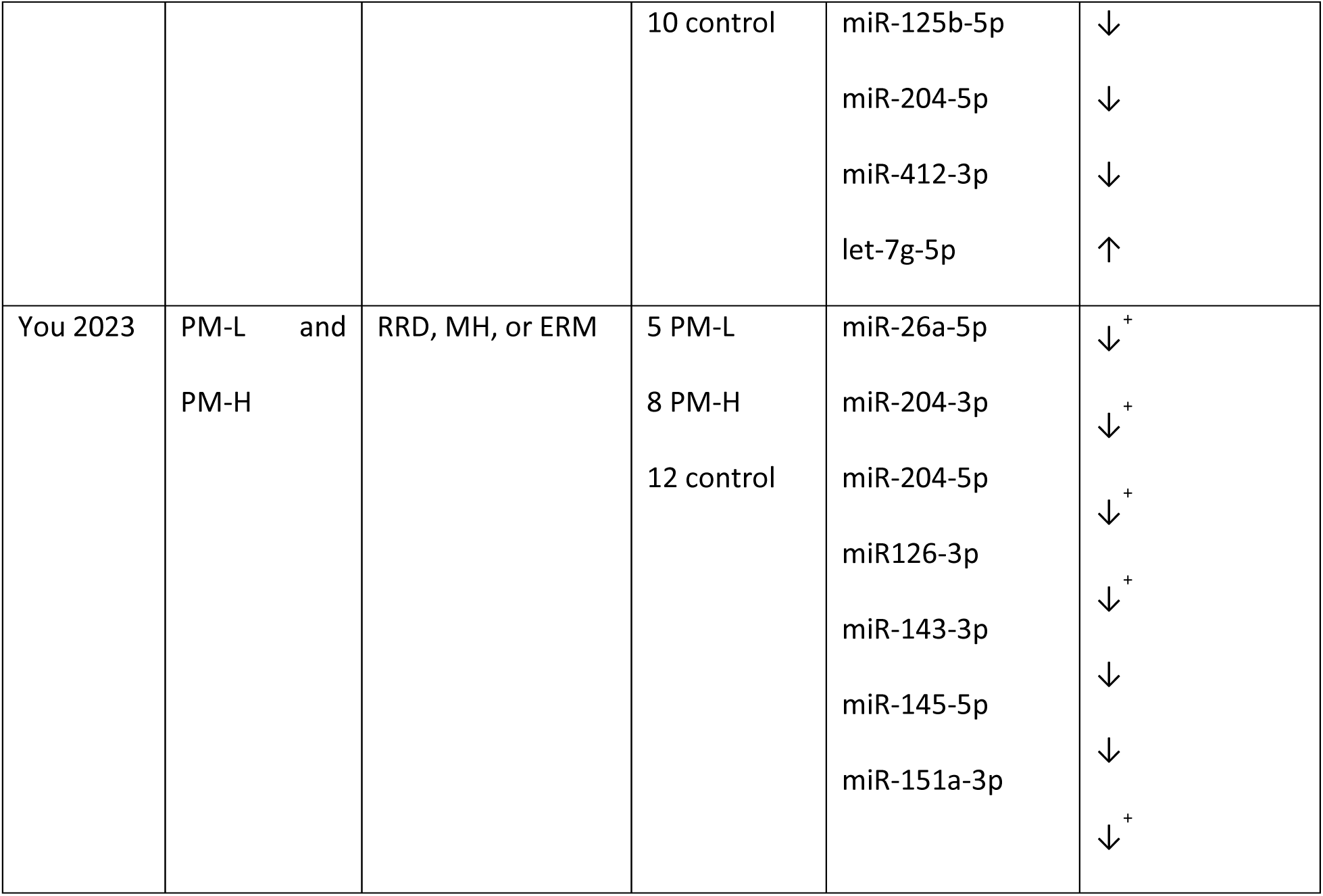
Dysregulated miRNA found within human vitreal exosomes. Summary of miRNA within vitreous humor exosomes that have thus far been studied and identified as dysregulated in comparison to the listed controls (p<0.05). UM: uveal melanoma, MH: macular hole, PDR: proliferative diabetic retinopathy, TRD: tractional retinal detachment, PM-L: low grade pathologic myopia, PM-H: high grade pathologic myopia, RRD: rhegmatogenous retinal detachment, ERM: epiretinal membrane. The table includes select miRNA that were emphasized in the primary study, see original publication for full list of dysregulated miRNAs. Up- or down- regulation noted with ‘+’ is comparing miRNA in PM-H to miRNA in PM-L, rather than to the control group.

| Study | Disease | Control | Sample Size | miRNA | Up- or Down-regulation |
| --- | --- | --- | --- | --- | --- |
| Ragusa<br>2015 | UM | Healthy cornea<br>donors | 6 UM<br>6 control | miR-21 | ↑ |
|  |  |  |  | miR-34a | ↑ |
|  |  |  |  | miR-146a | ↑ |
|  |  |  |  | miR-618 | ↑ |
| Liu 2022 | PDR | MH | 10 PDR<br>15 control | miR-9-3p | ↑ |
|  |  |  |  | miR-23b-3p | ↑ |
|  |  |  |  | miR-1285-3p | ↑ |
|  |  |  |  | miR-4685-3p | ↑ |
| Kot 2022 | PDR with<br>TRD | MH | 10 PDR with<br>TRD | miR-21-5p | ↑ |
|  |  |  |  | miR-125a-5p | ↓ |
|  |  |  | 10 control | miR-125b-5p<br>miR-204-5p<br>miR-412-3p<br>let-7g-5p | ↓<br>↓<br>↓<br>↑ |
| You 2023 | PM-L and<br>PM-H | RRD, MH, or ERM | 5 PM-L<br>8 PM-H<br>12 control | miR-26a-5p<br>miR-204-3p<br>miR-204-5p<br>miR126-3p<br>miR-143-3p<br>miR-145-5p<br>miR-151a-3p | ↓<sup>+</sup><br>↓<sup>+</sup><br>↓<sup>+</sup><br>↓<sup>+</sup><br>↓<br>↓<br>↓<sup>+</sup> |

Ragusa et al. was the first to look at exosomal miRNA in the vitreous humor, and they did so in the context of uveal melanoma (UM). The study looked at non-exosomal miRNA as well and found that there was a 90% overlap between non-exosomal and exosomal miRNA in UM, indicating that the non-exosomal miRNA may have altered by exosomes. Considering how exosomal miRNA reflect the parent cells, the study argues that miR-21 and miR-146a, which were found to be upregulated, come from neighboring neoplastic melanocytes. They show this by demonstrating that miR-146a was also upregulated in formalin-fixed paraffin-embedded UM specimens. A limitation of this study, however, is the choice of subjects. The vitreous humor was taken from enucleated eyes for the UM group and cornea donors for the control group. The miRNA composition, both exosomal and non-exosomal, could be altered by the removal of the eyes.

While shared miRNA dysregulation between Liu et al. and Kot et al. was discussed in the section on miRNA in PDR, the study by Liu et al. has further importance regarding exosomes. Liu et al. demonstrated that retinal Müller glia cells secrete exosomes containing miR-9-3p, and these miRNAs promote angiogenesis via overexpression of sphingosine 1-phosphate receptor 1 (S1P1). Again, this shows a direct connection between retinal cells and miRNA within vitreous humor exosomes. However, although this connection was demonstrated *in vivo*, it utilized a mouse model, which has limited transferability to humans. Further study in humans and animal models will be required to determine the exact role of exosomal miR-9-3p in PDR.

## Conclusions

The objective of this meta-analysis was to assess the potential usefulness of miRNAs identified in vitreous humor as biomarkers and diagnostic aids for a range of acquired retinal diseases. Among 126 initial studies surveyed, only 22 presented pertinent data concerning the composition of miRNAs in human vitreous biopsies within the context of retinal disease, covering conditions such as PDR, PVRD, PVRL, PM, AMD, and intraocular TB. Our revision of published data reveals promising prospects for utilizing miRNA molecular analysis in human vitreous biopsies as a feasible means of identifying and quantifying clinically relevant biomarkers. Moreover, the exploration of vitreous miRNAs holds potential for offering valuable insights into the molecular mechanisms underlying various retinal diseases. Nonetheless, significant challenges lie ahead, including the variability in extraction and purification methodologies, the establishment of robust criteria for selecting control samples, the identification of suitable and universally applicable housekeeping genes, and the validation of predictive capabilities for each proposed marker. Addressing these challenges will be crucial for advancing the clinical translation of miRNA-based diagnostics in retinal disease.

Our analysis revealed that variability across miRNA isolation and purification kits generally did not affect the consistency of results. However, differences in amplification methods may have contributed to contradictory findings. For instance, in three studies using three different kits (Kot et al. used miRCury, Friedrich et al. used TRizol, and Guo et al. used miRNeasy), all three found miR-142 to be upregulated in PDR (43, 60, 78). Nevertheless, while Kot et al. used qPCR for miRNA quantification, Liu et al. and Guo et al. initially used NGS, potentially accounting for their disparate findings regarding miR-9 dysregulation in PDR. Although no gold standard exists for miRNA quantification in vitreous biopsies, studies on limited volumes of serum and plasma recommend initial quantification with NGS followed by validation with qPCR to maximize miRNA detection sensitivity and reliability (95). While varying results among isolation kits and amplification techniques have been investigated in other human biofluids, the emerging nature of this field means there is yet no gold standard for miRNA extraction and quantification from vitreous humor, affecting the quality of results.

Due to the invasiveness of a vitrectomy, all subjects in the studies, including those considered ‘controls’, had a clinical indication for a vitrectomy, creating a lack of a *true control*. Most of the control cases required vitrectomies for idiopathic MH, ERM, or floaters, all of which are conditions caused by age-related changes to the vitreous. However, the pathophysiology of MH remains unknown, while the pathophysiology of ERM includes cell proliferation and fibrosis, which may also be occurring in the studied diseases, obscuring data (96, 97). For example, Russo et al. found that miR-19b, miR-24, and miR-142-3p were significantly upregulated in patients with idiopathic MH or ERM compared to idiopathic floaters (98). The miR-142 family was also found to be commonly upregulated in PDR when compared to controls with MH, indicating that there may be overlap in the pathogenesis of these idiopathic conditions and PDR.

For vitreous-derived miRNAs to be proven as biomarkers and indicators of pathogenesis, studies must use controls that do not have any underlying pathologies, but this is made challenging due to the invasiveness of a vitrectomy. Currently, most miRNA purification kits work reliably in fluid samples of ∼100-200 µl, while the human vitreous volume is estimated at ∼4.0 ml (99). A current alternative to vitrectomies would be to obtaining vitreous samples during intraocular injections. However, this restricts the collection of samples exclusively to those receiving the treatment in question. Future vitreous biopsy techniques with minimal invasiveness could involve the use of thinner and smaller catheters and needles, to extract a very small volume (<100 µl) from awake patients, eliminating the use of vitrectomy- or injection-dependent biopsies.

Variations in miRNA expression across studies could also be caused by different strategies for normalizing miRNA quantification data. Our studies reported 3 different normalization techniques – 7 used a housekeeping gene, 5 used an exogenous control, and 9 normalized against a weighted or unweighted average or median expression of the miRNA quantities. The housekeeping gene used by all 7 studies was the small nuclear-RNA U6, a widely used reference gene. A review of miRNA-analysis using qPCR in cancer studies showed that 84% of studies used U6 as the reference gene (100). However, the expression of U6 was shown to vary with experimental conditions and disease, suggesting potential limitations in its use as a universal housekeeping gene (101, 102). Other studies have shown that the ideal housekeeping gene varies based on disease state and the biological fluid being studied, and mathematical models such as geNorm, NormFinder, and BestKeeper should be used to identify a housekeeping gene that is optimal for vitreous humor (100, 103). This holds particular significance as a gold standard housekeeping gene would alleviate the challenge of small sample sizes in these studies, facilitating more straightforward pooling of data.

One possible solution to the variability in the expression of housekeeping genes is the use of an exogenous control. For example, cel-miR-39 -added to the vitreous samples-was used by several studies reviewed here. Exogenous controls studies in the serum of cancer patients indeed showed less inter-sample variability than endogenous controls (104). One drawback of exogenous controls, however, is that because they are added in just prior to RNA isolation, they do not control for experimental variability that occurs prior to this step (105). Other analysis strategies involve normalizing the expression of the target gene to the rest of the genes within a microarray, either by averaging them, finding the median, or identifying a few miRNAs that resemble the mean. While these strategies are mostly appropriate for high-throughput studies with several hundreds of miRNAs, they have been shown to produce more accurate results than reference genes in tumor samples (105, 106). One study, by Ando et al., used a housekeeping gene, as well as the median expression level, to normalize the quantification of the target genes(86). Until a standard housekeeping gene is identified and confirmed, normalization using a combination of housekeeping genes and general average values may be the best approach.

Another barrier to building on the reviewed studies to identify clinically applicable biomarkers is their validation, especially given their limited sample sizes. Two of the revised studies attempted to strengthen their findings by concurrently studying miRNA in the serum of their subjects. However, the two studies had opposing approaches. For example, Minezaki et al. found that miR-6513-3p, miR-138-2f-3p and miR-4445-3p were elevated in vitreous and serum of PVRL patients as compared to uveitis patients, but they did not compare the relative expression of miRNAs in vitreous vs serum for each patient (44). In contrast, Hirota et al. reported an increase in the expression of miR-23a, miR-320a, and miR-320b in the vitreous of PDR patients, as compared to the serum of the same PDR patients and as compared to vitreous samples from controls (40). This suggests that upregulation of these miRNAs in PDR is an intraocular-specific process, rather than a systemic one, providing new insights into PDR pathogenesis. The strategy used by Minezaki et al., looking for a positive correlation between vitreous and serum miRNA levels, may be more useful for validating biomarkers involved in systemic diseases. If successful, this approach can lead to serum biopsies replacing vitreous altogether, while still providing data that is clinically relevant for ocular and retinal diseases. The approach by Hirota et al. could be more useful for changes localized to the eye. Beyond biomarker validation, results that are tissue-dependent can carry with them potential mechanistic insights. Overall, there is currently little data to guide the interpretation of miRNA levels in vitreous vs serum, and how these comparisons should affect the potential of miRNA as diagnostic biomarkers.

Among the three most commonly dysregulated miRNAs in PDR, miR-142 emerged as consistently upregulated and specific to PDR, presenting itself as a potential biomarker. However, while KEGG analysis indicated pathway enrichments related to diabetes and diabetic retinopathy, further elucidation of miR-142’s role in the underlying pathology of PDR is necessary. On the other hand, miR-9 garnered substantial evidence supporting its involvement in the pathogenesis of PDR, thereby enhancing its potential as a biomarker. Notably, Liu et al. demonstrated elevated levels of miR-9 in exosomes derived from retinal Muller cells, with miR-9 facilitating angiogenesis through the targeting of the S1P1/AKT/VEGFR2 pathway. Moreover, miR-9’s angiogenic effects via S1P1 targeting in tumor endothelial cells underscore its significance (107). Additionally, KEGG analysis highlighted miR-9’s involvement in the p53 and MAPK pathways, both implicated in PDR pathogenesis (65, 66, 75). These findings collectively underscore miR-9’s potential as a crucial pathological indicator of PDR, notwithstanding its downregulation in the study by Kot et al., which could be attributed to population differences or methodological disparities in miRNA quantification.

Turning attention to PVRD, miR-21 emerged as dysregulated in four studies, with elevated levels correlating with PVRD pathogenesis. For instance, Usui-Ouchi et al. demonstrated TGF-β-induced upregulation of miR-21 expression in RPE cells, promoting RPE cell proliferation (81). Given the prominent role of TGF-β in inflammation and cell proliferation, and its targeting in PDR treatment, the study suggests a potential pathogenic role for miR-21 in diabetic retinopathy (108–110). Furthermore, miR-21-dependent promotion of fibrosis in various contexts, suggests its contribution to the retinal fibrosis observed in PVRD (111–113). Against the reported downregulation of miR-21 in the study by Toro et al., its consistent upregulation in three other studies suggests potential isoform-specific differences contributing to this discrepancy. Further confirmatory studies validating miR-21-5p upregulation in vitreous humor in PVRD would be valuable, given the established role of miR-21 in retinal and systemic fibrosis, cementing its emerging status as a pathological marker in PVRD.

In conclusion, it is important to acknowledge the limitations of the current analysis, notably the small sample sizes in most studies, and the variability in methodologies for miRNA purification, amplification, and quantification across studies. Additionally, the heterogeneous selection of control groups, ranging from healthy individuals to those with minimal risk of disease progression, poses a challenge to this emerging field. Nonetheless, our findings suggest that specific miRNA, including miR-142, miR-9, and miR-21, hold promise as diagnostic tools for their respective diseases. Further research is needed to validate and expand these findings and advance the clinical integration of miRNA as biomarkers for retinal diseases.

## Data Availability

All data produced in the present work are contained in the manuscript.

## List of Abbreviations

AMD: Age-related Macular Degeneration
ERM: Epiretinal Membrane
DR: Diabetic Retinopathy
MH: Macular Hole
miRNA: microRNA
NPDR: Non-Proliferative Diabetic Retinopathy
PDR: Proliferative Diabetic Retinopathy
PM: Pathological Myopia
PVR: Proliferative Vitreoretinopathy
PVRD: Proliferative vitreoretinal disease
PVRL: Primary Vitreoretinal Lymphoma
RD: Retinal Detachment
RRD: Rhegmatogenous Retinal Detachment

## Declarations

### Ethics approval and consent to participate

Not applicable

### Consent for publication

Not applicable

### Availability of data and materials

All data generated or analyzed during this study are included in this published article, and its supplementary information files.

### Authors contributions

DJ designed and conducted the research; collected and interpreted the data; wrote the manuscript. BG assisted in manuscript writing and editing. MT conceptualized the study and designed the research; assisted in data interpretation; wrote and edited the manuscript. All authors read and approved the final manuscript.

## Acknowledgements

Not applicable.

