## Supplemental Tables 1 and 2 for "Biomarker potential of vitreous microRNA in retinal disease: a meta-analysis"

**Supplemental Materials**

**Supplementary Table 1.** The 19 most commonly dysregulated (≥3 instances of dysregulation) miRNA species in proliferative diabetic retinopathy.

| **miRNA** | **Study** | **Arm** | **Up/Downregulation** |
|---|---|---|---|
| miR-142 | Kot 2022 | miR-142-3p | Up |
| | Friedrich 2020 | miR-142-3p | Up |
| | Kot 2022 | miR-142-5p | Up |
| | Guo 2021 | miR-142-5p | Up |
| miR-423 | Guo 2021 | miR-423-3p | Up |
| | Hirota 2014 | miR-423-5p | Up |
| | Smit-McBride 2020 | miR-423-5p | Up |
| | Guo 2021 | miR-423-5p | Up |
| miR-9 | Kot 2022 | miR-9-3p | Down |
| | Liu 2022 | miR-9-3p | Up |
| | Kot 2022 | miR-9-5p | Down |
| | Guo 2021 | miR-9-5p | Up |
| let-7a | Smit-McBride 2020 | let-7a | Up |
| | Guo 2021 | let-7a-3p | Up |
| | Guo 2021 | let-7a-5p | Up |
| let-7g | Smit-McBride 2020 | let-7g | Up |
| | Kot 2022 | let-7g-5p | Up |

| | | | |
|---|---|---|---|
| | Guo 2021 | let-7g-5p | Up |
| miR-1287 | Smit-McBride 2020 | miR-1287 | Down |
| | Guo 2021 | miR-1287-3p | Up |
| | Guo 2021 | miR-1287-5p | Up |
| miR-15a | Hirota 2014 | miR-15a | Up |
| | Kot 2022 | miR-15a-5p | Up |
| | Guo 2021 | miR-15a-5p | Up |
| miR-16 | Smit-McBride 2020 | miR-16 | Up |
| | Kot 2022 | miR-16-5p | Up |
| | Guo 2021 | miR-16-5p | Up |
| miR-181a | Guo 2021 | miR-181a-2-3p | Up |
| | Guo 2021 | miR-181a-3p | Up |
| | Guo 2021 | miR-181a-5p | Up |
| miR-185 | Smit-McBride 2020 | miR-185 | Up |
| | Friedrich 2020 | miR-185-5p | Up |
| | Guo 2021 | miR-185-5p | Up |
| miR-30d | Guo 2021 | miR-30d-3p | Up |
| | Guo 2021 | miR-30d-5p | Up |
| | Smit-McBride 2020 | miR-30d-star | Down |
| miR-320a | Hirota 2014 | miR-320a | Up |
| | Smit-McBride 2020 | miR-320a | Up |
| | Guo 2021 | miR-320a-3p | Up |
| miR-320b | Hirota 2014 | miR-320b | Up |

| | | | |
|---|---|---|---|
| | Smit-McBride 2020 | miR-320b | Up |
| | Guo 2021 | miR-320b | Up |
| miR-326 | Friedrich 2020 | miR-326 | Up |
| | Guo 2021 | miR-326 | Up |
| | Friedrich 2020 | miR-326-5p | Up |
| miR-425 | Guo 2021 | miR-425-3p | Up |
| | Guo 2021 | miR-425-5p | Up |
| | Smit-McBride 2020 | miR-425-star | Up |
| miR-486 | Guo 2021 | miR-486-3p | Up |
| | Smit-McBride 2020 | miR-486-5p | Up |
| | Guo 2021 | miR-486-5p | Up |
| miR-92a | Smit-McBride 2020 | miR-92a | Up |
| | Kot 2022 | miR-92a-3p | Up |
| | Guo 2021 | miR-92a-3p | Up |
| miR-93 | Hirota 2014 | miR-93 | Up |
| | Mammadzada 2019 | miR-93 | Up |
| | Guo 2021 | miR-93-5p | Up |
| miR-223 | Guo 2021 | miR-223-3p | Up |
| | Guo 2021 | miR-223-5p | Up |
| | Smit-McBride 2020 | miR-223-star | Down |

**Supplementary Table 2.** The 25 most commonly dysregulated (≥3 instances of dysregulation) miRNA species in proliferative vitreoretinal disease.

| **miRNA** | **Study** | **Disease*** | **Arm** | **Up/Downregulation** |
|---|---|---|---|---|
| miR-9 | Kot 2022 | PDR | miR-9-3p | Down |
| | Liu 2022 | PDR | miR-9-3p | Up |
| | Kot 2022 | PDR | miR-9-5p | Down |
| | Guo 2021 | PdR | miR-9-5p | Up |
| | Toro 2020 | RD with PVR | miR-9-5p | |
| miR-423 | Guo 2021 | PDR | miR-423-3p | Up |
| | Hirota 2014 | PDR | miR-423-5p | Up |
| | Smit-McBride 2020 | PDR | miR-423-5p | Up |
| | Guo 2021 | PDR | miR-423-5p | Up |
| | Usui-Ouchi 2016 | PVRD | miR-423-5p | Up |
| miR-139 | Smit-McBride 2020 | PDR | miR-139-3p | Up |
| | Guo 2021 | PDR | miR-139-3p | Up |
| | Usui-Ouchi 2016 | PVRD | miR-139-5p | Down |
| | Toro 2020 | RD with PVR | miR-139-5p | Up |
| miR-142 | Kot 2022 | PDR | miR-142-3p | Up |
| | Friedrich 2020 | PDR | miR-142-3p | Up |
| | Kot 2022 | PDR | miR-142-5p | Up |
| | Guo 2021 | PDR | miR-142-5p | Up |
| | Smit-McBride 2020 | PDR | miR-16 | Up |
| | Usui-Ouchi 2016 | PVRD | miR-16 | Up |

| | | | | |
|---|---|---|---|---|
| | Kot 2022 | PDR | miR-16-5p | Up |
| | Guo 2021 | PDR | miR-16-5p | Up |
| miR-21 | Usui-Ouchi 2016 | PVRD | miR-21 | Up |
| | Toro 2020 | RD with PVR | miR-21-3p | Down |
| | Kot 2022 | PDR | miR-21-5p | Up |
| | Guo 2021 | PDR | miR-21-5p | Up |
| miR-223 | Guo 2021 | PDR | miR-223-3p | Up |
| | Guo 2021 | PDR | miR-223-5p | Up |
| | Toro 2020 | RD with PVR | miR-223-5p | Down |
| | Smit-McBride 2020 | PDR | miR-223-star | Down |
| miR-486 | Guo 2021 | PDR | miR-486-3p | Up |
| | Toro 2020 | RD with PVR | miR-486-3p | Up |
| | Smit-McBride 2020 | PDR | miR-486-5p | Up |
| | Guo 2021 | PDR | miR-486-5p | Up |
| miR-92a | Smit-McBride 2020 | PDR | miR-92a | Up |
| | Usui-Ouchi 2016 | PVRD | miR-92a | Up |
| | Kot 2022 | PDR | miR-92a-3p | Up |
| | Guo 2021 | PDR | miR-92a-3p | Up |
| miR-204 | Usui-Ouchi 2016 | PVRD | miR-204 | Down |
| | Guo 2021 | PDR | miR-204-3p | Up |
| | Kot 2022 | PDR | miR-204-5p | Down |
| miR-27a | Mammadzada 2019 | PDR | miR-27a | Up |
| | Guo 2021 | PDR | miR-27a-3p | Up |

| | | | | |
|---|---|---|---|---|
| | Toro 2020 | RD with PVR | miR-27a-5p | Down |
| miR-361 | Kot 2022 | PDR | miR-361-3p | Down |
| | Guo 2021 | PDR | miR-361-5p | Up |
| | Toro 2020 | RD with PVR | miR-361-5p | Up |
| let-7a | Smit-McBride 2020 | PDR | let-7a | Up |
| | Guo 2021 | PDR | let-7a-3p | Up |
| | Guo 2021 | PDR | let-7a-5p | Up |
| let-7g | Smit-McBride 2020 | PDR | let-7g | Up |
| | Kot 2022 | PDR | let-7g-5p | Up |
| | Guo 2021 | PDR | let-7g-5p | Up |
| miR-1287 | Smit-McBride 2020 | PDR | miR-1287 | Down |
| | Guo 2021 | PDR | miR-1287-3p | Up |
| | Guo 2021 | PDR | miR-1287-5p | Up |
| miR-15a | Hirota 2014 | PDR | miR-15a | Up |
| | Kot 2022 | PDR | miR-15a-5p | Up |
| | Guo 2021 | PDR | miR-15a-5p | Up |
| miR-16 | Smit-McBride 2020 | PDR | miR-16 | Up |
| | Kot 2022 | PDR | miR-16-5p | Up |
| | Guo 2021 | PDR | miR-16-5p | Up |
| miR-181a | Guo 2021 | PDR | miR-181a-2-3p | Up |
| | Guo 2021 | PDR | miR-181a-3p | Up |
| | Guo 2021 | PDR | miR-181a-5p | Up |

| | | | | |
|---|---|---|---|---|
| miR-185 | Smit-McBride 2020 | PDR | miR-185 | Up |
| | Friedrich 2020 | PDR | miR-185-5p | Up |
| | Guo 2021 | PDR | miR-185-5p | Up |
| miR-30d | Guo 2021 | PDR | miR-30d-3p | Up |
| | Guo 2021 | PDR | miR-30d-5p | Up |
| | Smit-McBride 2020 | PDR | miR-30d-star | Down |
| miR-320a | Hirota 2014 | PDR | miR-320a | Up |
| | Smit-McBride 2020 | PDR | miR-320a | Up |
| | Guo 2021 | PDR | miR-320a-3p | Up |
| miR-320b | Hirota 2014 | PDR | miR-320b | Up |
| | Smit-McBride 2020 | PDR | miR-320b | Up |
| | Guo 2021 | PDR | miR-320b | Up |
| miR-326 | Friedrich 2020 | PDR | miR-326 | Up |
| | Guo 2021 | PDR | miR-326 | Up |
| | Friedrich 2020 | PDR | miR-326-5p | Up |
| miR-425 | Guo 2021 | PDR | miR-425-3p | Up |
| | Guo 2021 | PDR | miR-425-5p | Up |
| | Smit-McBride 2020 | PDR | miR-425-star | Up |
| miR-93 | Hirota 2014 | PDR | miR-93 | Up |
| | Mammadzada 2019 | PDR | miR-93 | Up |
| | Guo 2021 | PDR | miR-93-5p | Up |

*PDR = proliferative diabetic retinopathy, RD = retinal detachment, PVR = proliferative vitreoretinopathy, PVRD = proliferative vitreoretinal disease

14
